# Mathematical Modeling of Coronavirus Reproduction Rate with Policy and Behavioral Effects

**DOI:** 10.1101/2020.06.16.20133330

**Authors:** Rabeya Anzum, Md. Zahidul Islam

## Abstract

In this paper a modified mathematical model based on the SIR model used which can predict the spreading of the corona virus disease (COVID-19) and its effects on people in the days ahead. This model considers all the death, infected and recovered characteristics of this disease. To determine the extent of the risk posed by this novel coronavirus; the transmission rate (*R*_0_) is utilized for a time period from the beginning of spreading virus. Particularly it includes a novel policy to capture the Ro response in the virus spreading over time. The model estimates the vulnerability of the pandemic according to John H. Cochrane’s method with a prediction of new cases by estimating a time-varying *R*_0_ to capture changes in the behavior of SIR model implies to new policy taken at different times and different locations of the world. This modified SIR model with the different values of *R*_0_ can be applied to different country scenario using the real time data report provided by the authorities during this pandemic. The effective evaluation of *R*_0_ can forecast the necessity of lockdown as well as reopening the economy.

## 1. Introduction

Mathematical modeling of the any disease spreading is a very important tool to predict the possible number of infected people as well as the spreading scenario.[1], [2], [3], [4]. As every disease have its own specific biological characteristics so to tackle real life situations, the established models need to be modified with respect to the specific cases.[5], [6].In December 2019, Coronavirus disease 2019 (COVID-19) emerged in China and then rapidly spread all over the world [7]. The World Health Organization (WHO) renamed the epidemic disease caused by 2019-nCoV as strain severe acute respiratory syndrome coronavirus 2 (SARS-CoV-2) on 11 February 2020 [8], [9].This is a new virus and the world is facing a new situation [10] as there is no vaccine has made to combat this virus. In this situation, on 30 January 2020, World Health Organization (WHO) declared it to be a Public Health Emergency of Global Concern [11]. As of 20 May 2020, the disease was confirmed in more than 4998097 cases reported globally and there are 325304 death cases reported. World health Organisation (WHO) declared it a pandemic situation caused by coronavirus spreading [12].

In this paper, the literature review consists of some relevant mathematical models which tried to describe the dynamics of the evolution of COVID-19. Some of the phenomenological models tried to generate and assess short-term forecasts using the cumulative number based on reported cases. The SIR model is a traditional one to predict the vulnerability of a pandemic and also can be used to predict the future scenario of coronavirus cases. However, this model is modified [13], [14] including other variables to calibrate the possible infection rate with time being. Coronavirus transmission (how quickly the disease spreads) is indicated by its reproduction number (*R*_0_, pronounced as R-nought or r-zero), which indicates the actual number of people to whom the virus can be transmitted by a single infected case. WHO predicted (on Jan. 23) *R*_0_, would be between 1.4 and 2.5. Other research measured Ro with various values somewhere between 3.6 and 4.0, and 2.24. The long-used value of Ro is 1.3 for flu and 2.0 for SARS [15][2]. In this research, firstly SIR model is simulated taking a constant value of *R*_0_ 0.5 to observe the response. Since reducing the face to face contact among people and staying home in lockdown can improve in reducing the further infection rate, *R*_0_ is taken as a time varying constant rather than a fixed one to observe the overall scenario of coronavirus spreading.

The organization of the paper is as follows: A brief study of some mathematical models correlated to coronavirus spreading is extracted in section 2, literature review part, section 3 contains the Modeling of coronavirus spreading. Section 4 is the result and discussion part and section 5 concludes the paper.

## 2. Literature Review

The SIR epidemic model which is used in this work is one of the simplest compartmental models firstly used by Kermack and McKendrick (1927).Compartmental model denotes mathematical modeling of infectious diseases where the population is separated in various compartments for example, S, I, or R, (Susceptible, Infectious, or Recovered). Many works have been undergone during the coronavirus pandemics utilizing the compartmental models [16], [17] and Imperial College COVID-19 Response Team (2020) provides a useful overview of this classic model to show that these models can be applied to understand the current health hazards. Much interest has been generated among economists to identify the impact of current pandemic on economic sectors by exploring compartmental models along with standard economic models using econometric techniques. [18], [19].

It was argued by the economists that many of the parameters controlling the move among compartments are not structural however it depends on individual decisions and policies. For example, according to Eichenbaum et al. [20] and Farboodi, Jarosch and Shimer [21], the number of new infections is a function of the endogenous labor supply and consumption choices of individuals which is determined by the rate of contact and by the standard decision theory models where the rate of contact is amenable. Similarly, the death rates and recovery rates are not just clinical parameters. Moreover, it can be used as functions of policy decisions for example by expanding emergency hospital capacity may increase the recovery rate and decrease death rates. Also, in this case the fatality ratio is a complex function because it depends on many clinical factors. Therefore, the selection-into-disease mechanisms are themselves partly the product of endogenous choices [22].

Moreover, concerning with the identification problems of compartmental models economists Atkeson [23] and Korolev [24] have found that these models lack many set of parameters that is able to fit the observed data efficiently before the long-run degradable consequences. Some of the researchers Linton [25] and Kucinskas [26] used time-series models utilizing the econometric tradition other than using compartmental models.

However, many of the economists are pushing the study of compartmental models in a multitude of dimensions. Acemoglu et al. [27] and Alvarez et al. [28] identified that the optimal lockdown policy for a planner who wants to control the fatalities of a pandemic while minimizing the output cost of the lockdown. Berger et al. [29] examine the role of testing and case-dependent quarantines. Bethune and Korinek [30] estimate the infection externalities associated with COVID-19. Bodenstein et al. [31] examine a compartmental model with a multi-sector dynamic general equilibrium model. Other researchers like Garriga, Manuelli and Sanghi [32],Hornstein [33], and Karin et al. [34] study a variety of containment policies. Toda [35], estimated a SIRD model to explores the optimal mitigation policy that controls the timing and intensity of social distancing. Flavio Toxvaerd[36] also developed a simple economic model emphasizing on endogenous social distancing.

Furthermore, many of the economists commented that coronavirus infection transmission cannot be a biologically induced constant rather it can be varied with human behavior and the change in human behavior can be predicted in response to changing social policies. Another form of multi-risk SIR Model with assumed a targeted lockdown period provided by the economists Daron Acemoglu et al [37] which is an epidemiology models with economic incentives. The authors of this paper argued about the herd immunity that might be much lower if the super spreaders like people in hospitals, emergency service providers, bus drivers, etc are set to immune first.

Therefore, in this paper we use John H. Cochrane’s methodology by allowing the infection rates changeable over time by imposing social distancing among people. Moreover, we tried to focus on the reproduction rate *R*_0_ which is not a constant but varies with time based on the demographic and policy heterogeneity.

## 3. Mathematical SIR Model

According to the conventional SIR model, for the N number of constant populations, each of whom may be in one of five states with respect to time implies that:

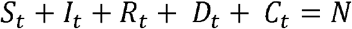

Where the parameters denote:

S = Susceptible,

I = Infected,

R = Resolving,

D = Number of dead from pandemic,

C = Number of recovered people,

N = Total number of population.

While a susceptible person can be affected by the disease when comes in contact with an infectious person.

### A. Modified-SIR model

The SIR model is used to predict the vulnerability of any type pandemic which may not be applicable to coronavirus cases since this model assumes the reproduction rate as a constant. The virus seems to be diminished when all affected people will be recovered which is practically not possible. The affected people by corona virus are highly contagious to other people who come in contact to them. Thus, the spreading of coronavirus infection is increasing day by day. Therefore, a mathematical model of the spreading of it can help predicting its vulnerability and while to take some efficient measures to lowering the contact rate.

The modified behavioral SIR model is presented by [38]:

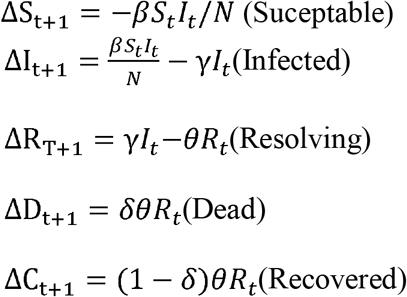

Where, β= Number of contacts per day. Again, lowering the rate of further reproduction does not stop the pandemic immediately. The modified version of SIR model shows exponential decays of reproduction rate other than fixed timeframe to be diminished. A person who can be affected meets β number of people in a day. Therefore, 1/N of the total population is becoming infected by coronavirus each day, so that *βS* _*t*_ *I* _*t*_ */N* is the number of infected people each day.The parameters varies over time to capture behavioral or policy changes such as social distancing. Therefore, the modified SIR model can be parameterized in terms of the *R*_0_ =reproduction rate.

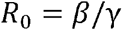

The number of infection spreading from infected person = number of possible contacts (per day) × the number of days the affected people remain infectious.

In the SIR model, β is a constant and hence we use a constant *R*_0_ =reproduction rate. Eventually the exponential growth of the disease then the growth can be confined by the reducing the number of susceptible people in the total number of population. Moreover, each of the infected people is considered to be recovered people after certain time. But in actual scenario it is not happening because of the death from coronavirus infection. In the actual SIR model lowered β was used. However, by lowering the reproduction rate *R*_0_, the further spread of coronavirus can be suppressed.

### B. Cochrane’s Behavioral SIR Model [39]

After the lockdown most of the people reduced their contacts which decrease the proportionally of the increased number of infectious people. Since β is the rate of susceptible person so that a logarithmic function can be used as it can’t be negative.

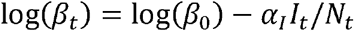

However, due to absence of sufficient continuous testing the number of infectious people at any time cannot be predicted. Therefore, the further expansion model can be expressed as utilizing the current death rate:

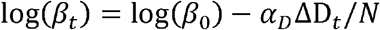

The logarithmic function was used to get the idea in the early declines while super spreading factor activities are eliminated.

### C. Parameter estimation according to Cochrane’s method

Let us assume the total number of populations is 1 million. In this model we used the same parameter value as used by the researcher Chad and Jesús [38]. Hereby,

γ =0.2 which is used for 5 days of remain infectiousness in an average,

θ=0.1 implies that 10 days in an average to get infected by coronavirus for which it may cause death before it resolves.

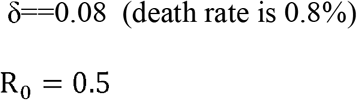

Which is used in modified SIR model such that,

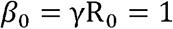

As per modified SIR model 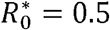 and*β*^*^ 0.1

To estimate α, β=*β*^*^.

For 50 daily deaths per million and assumingR_0_ = 0.5, α_*D*_ can be solved using the following:

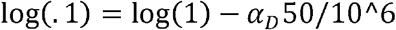

If the death rate is 1%, therefore the calibration of α is calculated at an infection rate of 0.5%. i.e. 5000 death at a million population.

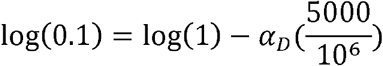

**Figure 1:**
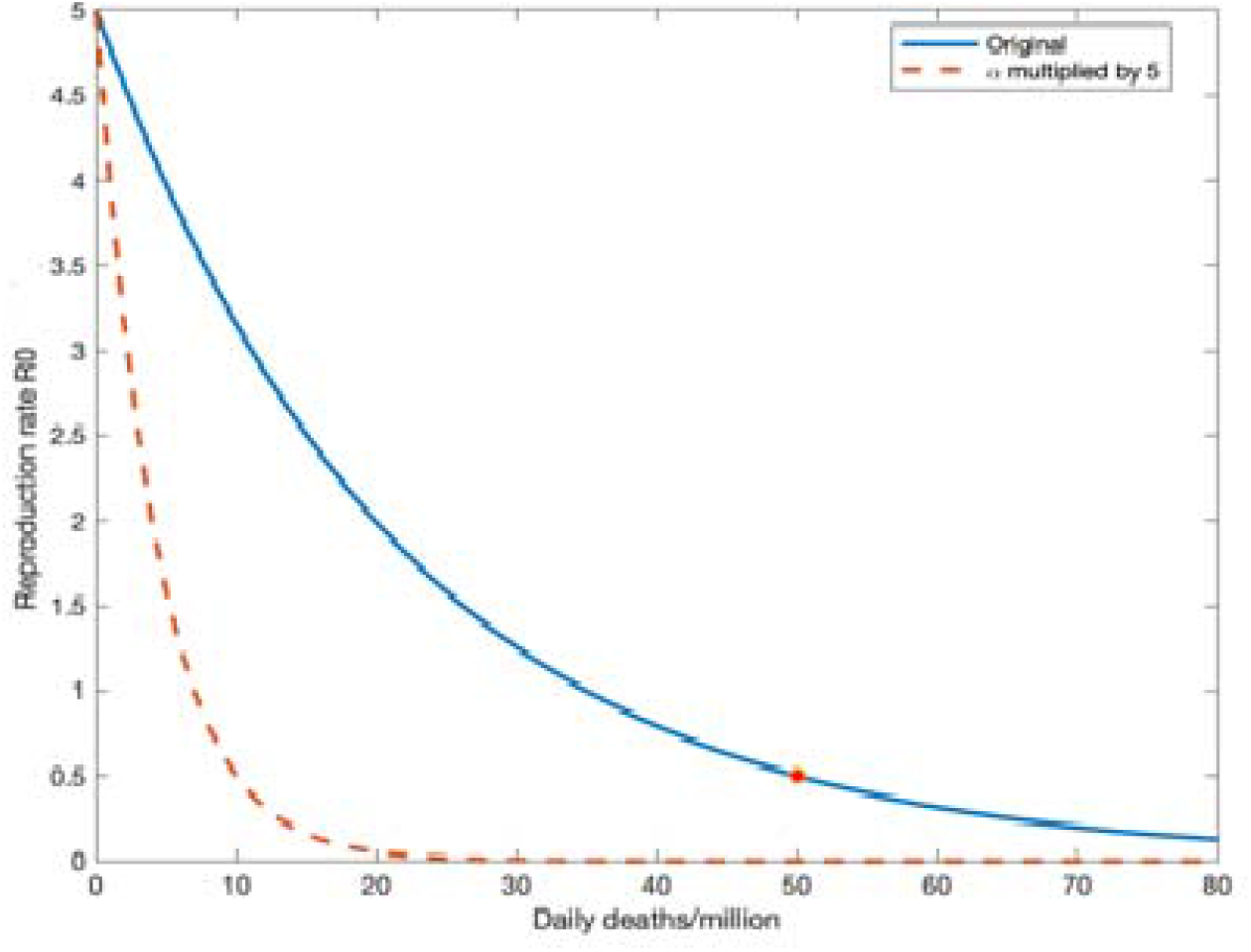
Reproductin rate, R_0_ calibration [39]

Here the reproduction rate R_0_ is assumed as a function of number of deaths calculated for one million population size. The red point in the graph indicates the calibration point while 50 death case is found on daily basis and R_0_ 0.5. The calibration response showed in dashed line (red) is the most acute response, which can be elaborated by utilizing different R_0_ values.

## 4. Results and Analysis of Cochrane’s Method

The standard SIR model simulation with those above-mentioned parameters values is as follow:

**Figure 2:**
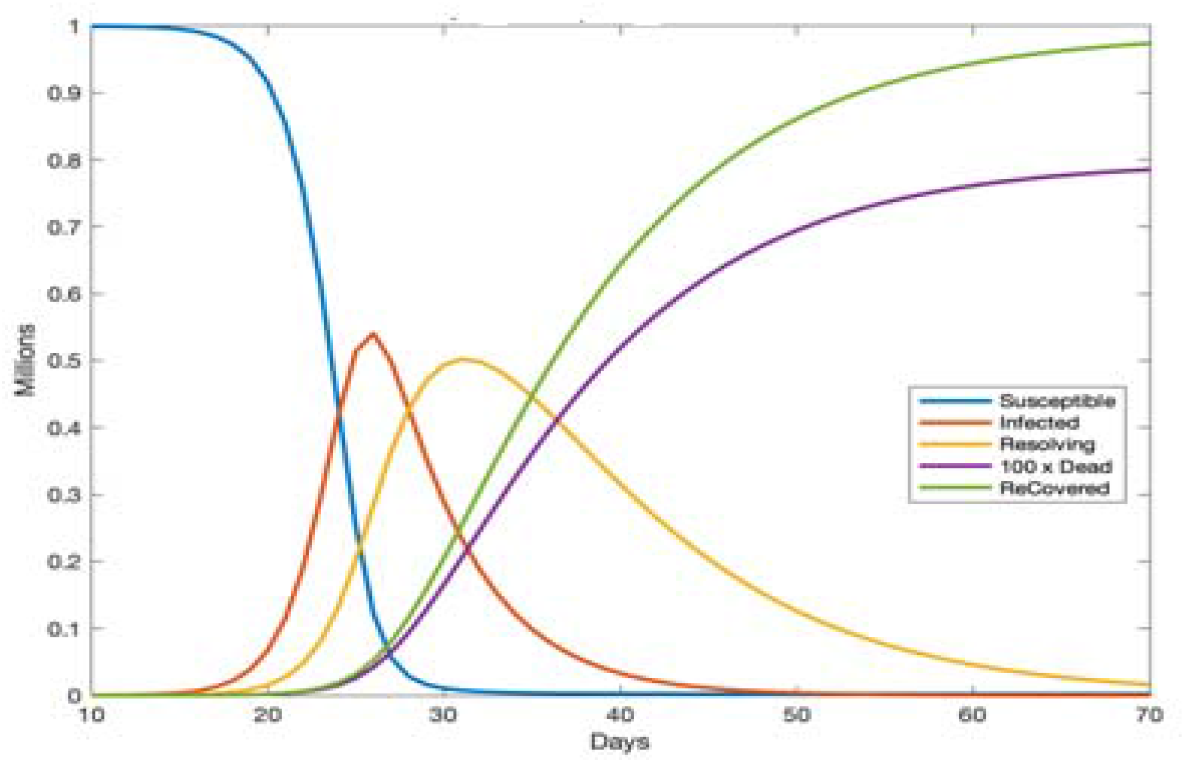
SIR model using R_0_ = 0.5 [39]

From the graph, the first day starts with only one infected person then the infection rate in the graph increases exponentially with the increasing number of new infected people while it shows the pick value at almost half the population. However, after 25 days of the infection spreading the herd immunity begun while the maximum number of infected people was found. The number of sick people who are resolving peaks after few days. The pandemic period is over 2 months and it is expected to go away after that. While everyone of the total 1 million populations get infected and that results into 0.8% or 8000 death.

Furthermore, the behavioral modified SIR model simulation, the system reacts by reducing the transmission rate meaning that the varied values of R_0_ function.

**Figure 3:**
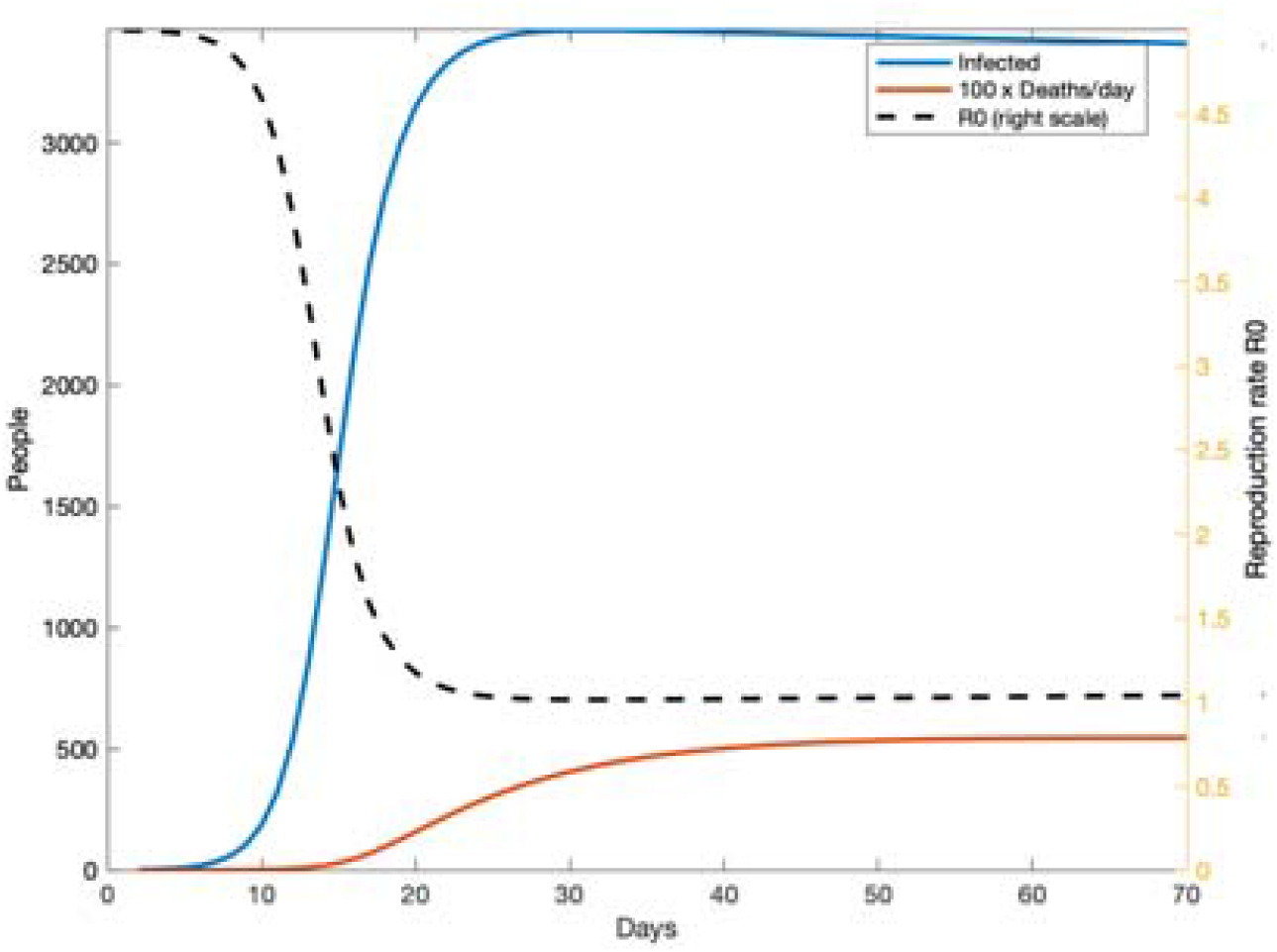
Behavioral SIR model with varied R_0_ [39]

The graph shows that about 4000 people will be infected but not the total 1 million populations. The pandemic gets going exponentially (blue line), while the infection rate reach 1000 per million there is a noticeable acute reduction in the further reproduction rate indicated in black colored dashed line.

Moreover, the transmission rate R_0_ is asymptotes in this model, with a decline in the number of infection rate as well as less number of death cases per day. In this process, it may take a long period to achieve herd immunity if all the initiatives taken in order to lessening contacts are stopped. Assuming the reproduction rate is asymptotes to R_0_ = 1, the following graph shows:

**Figure 4:**
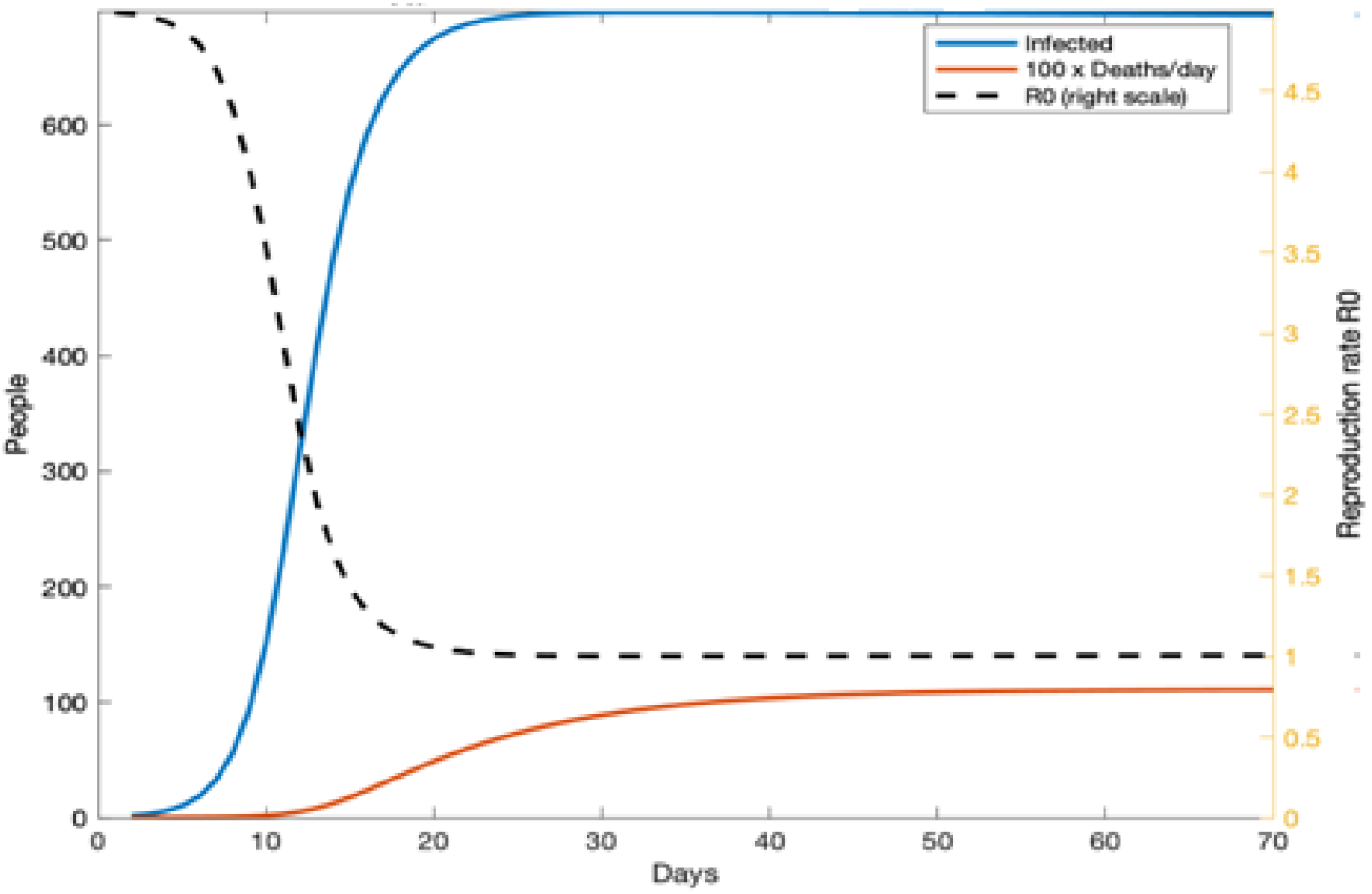
Behavioral SIR model with Varied R_0_ = 1 [39]

This simulation graph shows an aggressive response, α is increased by a factor of 5 showed in the dashed line (red) in the first graph. In this graph from the vertical scale it is clear that, the less people are vulnerable to the infections implies that the overall rate of infection is comparatively low. Being stricter about the social gathering and possible contacts the situation doesn’t change since the reproduction or transmission rate is still asymptotes, such that R_0_ declines to one. However, in such scenario, we get low infection rate on daily basis. After reducing the number of infected people over time, letting R_0_ varies over time and α is increasing by a factor of 2 for 100 days the following graph shows:

**Figure 5:**
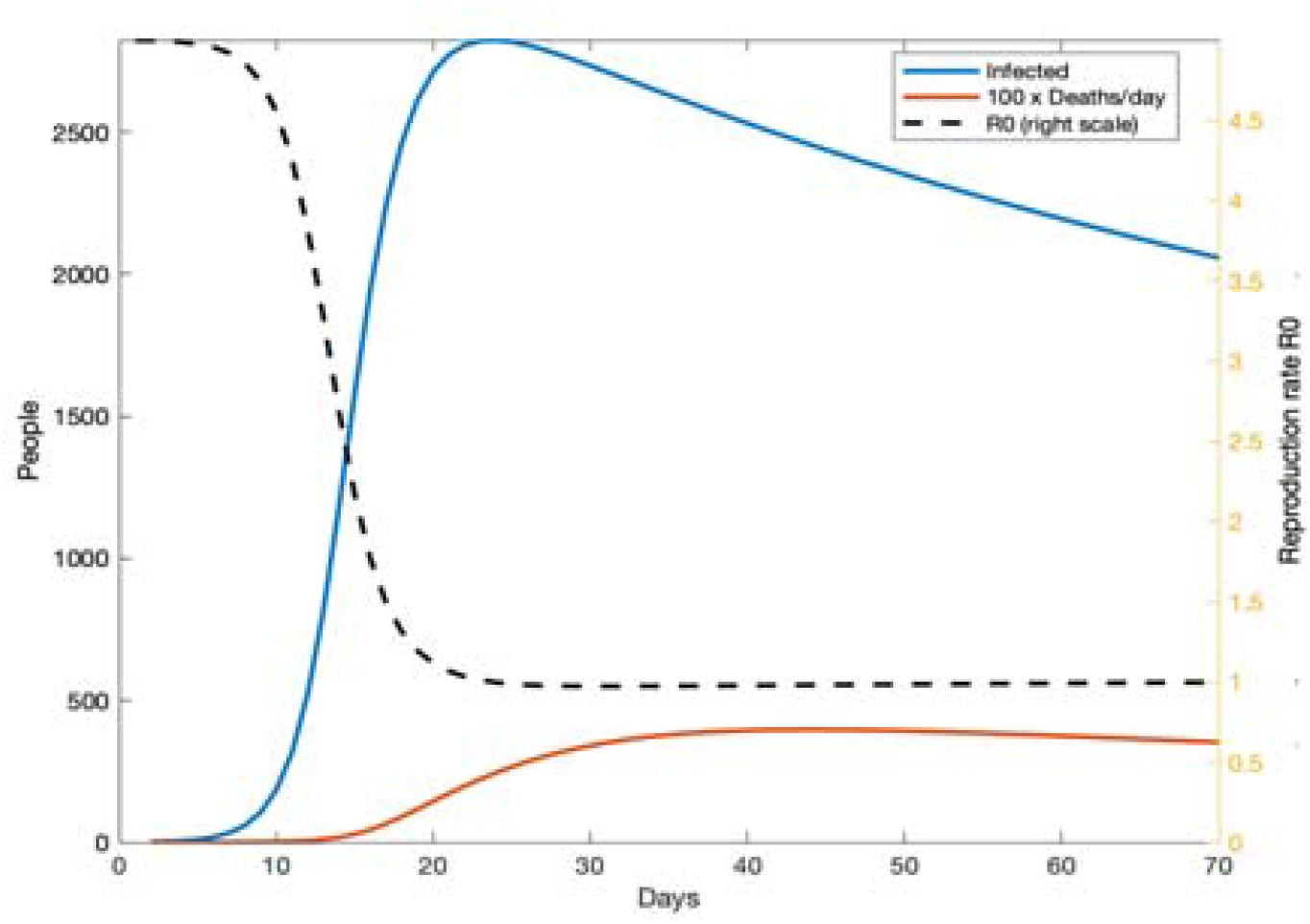
Behavioral SIR model with α increasing [39]

Therefore, the simulation of modified SIR model (when people care about the death rate rather than the reproduction of infection rate) gives the following graph:

**Figure 6:**
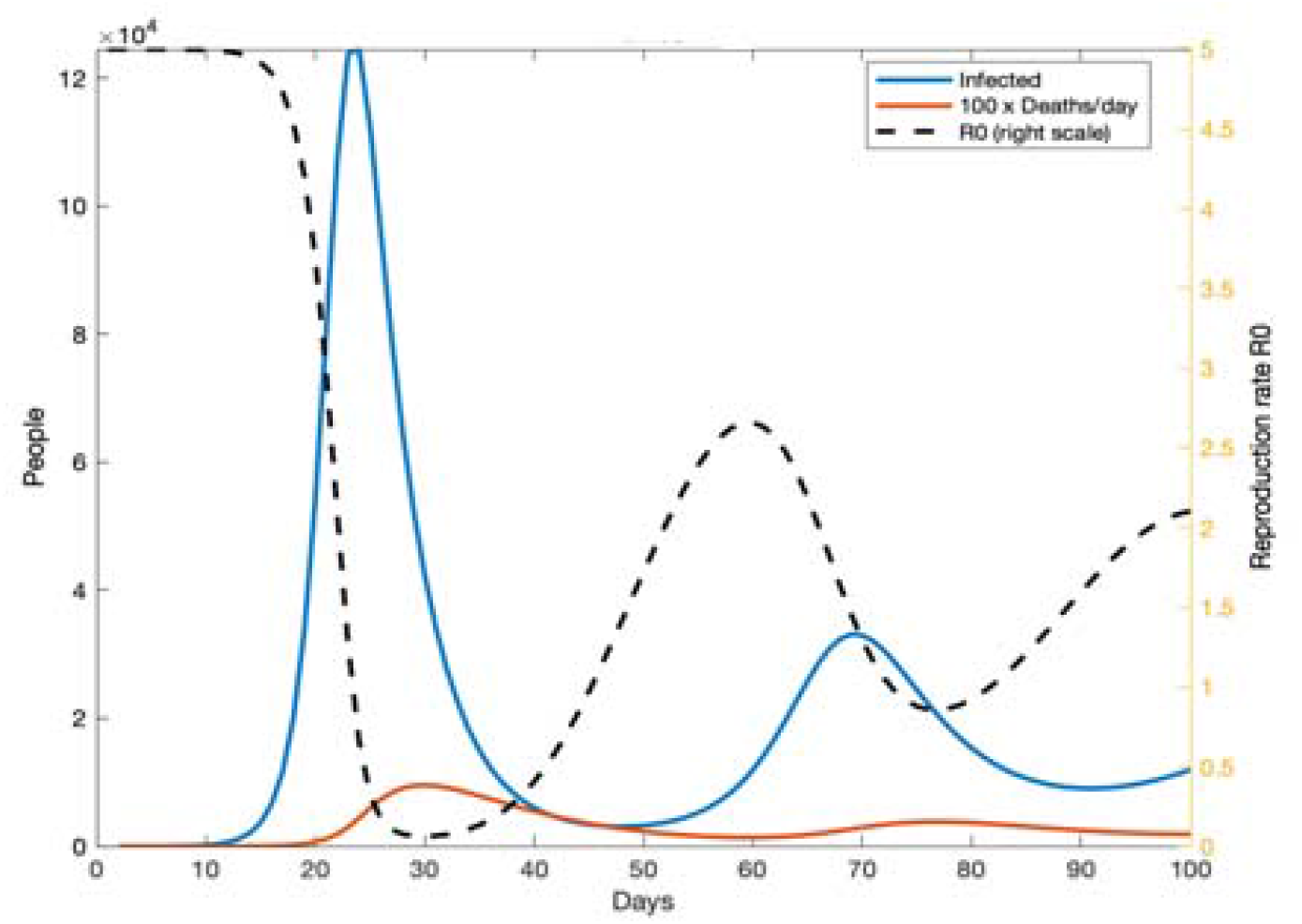
Modified SIR model with behavior and policy response [39]

Since after a few weeks’ deaths occur by the infections is found steady over time. Then the situation is considered under control. After letting people go back to the normal life with no restriction, the situation may get back with more severe affects. Then the coronavirus pandemic might be uncontrolled with high deaths in the first wave while by taken possible measures and consciousness of people to avoid contact can reduce the infection rate and death rate. Furthermore, after lowering the death rate people ease up and a second wave of infection rate appears so forth. Therefore, the reproduction rate R_0_ cannot be constant as it varies with the behavioral change of people and policy taken to control the pandemic.

## 5. Conclusion

The COVID-19 is a current global issue which spread in almost every country of the world and caused restriction to the free movement of people resulting a massive economical loss worldwide. Transmission rate (R_0_) is the number of newly infected individuals derived from a single case is the factor that can calibrated the reproduction rate of coronavirus utilizing the SIR model. In the traditional SIR model, taking Ro as a constant cannot predict the actual scenario of coronavirus spreading. The value of R_0_ can be different for different places and time period. Moreover, Ro can be changed with the behavioral changes of people because of the adopted policies by respective authorities. Therefore, in this research R_0_ is not considered a constant rather it is used as a time varying function. By taken the possible measures to reduce the social contact R_0_can be minimized with the time causing less death and forecasting to reopen the economy.

## Data Availability

All data referred to in the manuscript are available in Google scholar and google.

